# Escherichia marmotae - a human pathogen easily misidentified as Escherichia coli

**DOI:** 10.1101/2022.01.26.22269802

**Authors:** Audun Sivertsen, Ruben Dyrhovden, Marit Gjerde Tellevik, Torbjørn Sæle Bruvold, Eirik Nybakken, Dag Harald Skutlaberg, Ingerid Skarstein, Øyvind Kommedal

**Affiliations:** Department of Microbiology, Haukeland University Hospital, Bergen, Norway; University of Bergen, Bergen, Norway

## Abstract

We hereby present the first descriptions of human invasive infections caused by *Escherchia marmotae*, a recently described species that encompasses the former *“Escherichia* cryptic clade V”. We report four cases, one acute sepsis of unknown origin, one post-operative sepsis after cholecystectomy, one spondylodiscitis and one upper urinary tract infection. Cases were identified through unsystematic queries in a single clinical lab during six months. Through genome sequencing of the causative strains combined with available genomes from elsewhere we demonstrate *E. marmotae* to be a likely ubiquitous species containing genotypic virulence traits associated with *Escherichia* pathogenicity. The invasive isolates were scattered among isolates from a range of non-human sources, thus indicating inherent virulence in multiple phylogenetic lineages. Pan genome analyses indicate that *E. marmotae* has a large accessory genome and is likely to obtain ecologically advantageous traits like genes encoding antimicrobial resistance. Reliable identification might be possible by MALDI-ToF MS, but relevant spectra are missing in commercial databases. *E. marmotae* can be identified through 16S rRNA sequencing. *Escherichia marmotae* could represent a relatively common human pathogen and improved diagnostics will provide a better understanding of its clinical importance.

## Introduction

The type strain of *Escherichia marmotae* was isolated from the gut of the himalayan marmot (*Marmota himalayana*) and described as a novel species in 2015 (1). It encompasses the environmental *Escherichia* species previously referred to as “*Escherichia* cryptic clade 5”. Recently, *E. marmotae* was predicted to represent a potential human pathogen by *in vitro* infection assays and virulence gene characterisation (2). The presence of virulence factors may be linked to virulent *E. coli* occupying the same environmental niche in the vertebrate gut and it was also suggested that the *astA* gene encoding the heat-stable enterotoxin 1 in entero-aggregative *E. coli* originates from *E. marmotae* (3). *However, until now E. marmotae* has not been reported from human non-enteric samples and historically the “*Escherichia* cryptic clade 5” has been considered an environmental species (4, 5), a rare finding in human enteric samples and of low pathogenic potential (6).

Here, we present four case reports where *E. marmotae* was isolated as the likely pathogen of invasive human infections. The cases represent epidemiologically unrelated community acquired infections and were all discovered in 2021 over a period of six months. We also characterize the isolates using whole genome sequencing and investigate the phylogenetic relationships between our human invasive strains and the 36 *E. marmotae* strains with complete or draft whole genomes available in NCBI GenBank, as well as other strains representing the diversity within the *Escherichia* genus.

*E. coli* is among the most important human pathogens and the most common cause of Gram-negative bloodstream infections in adult patients. The finding that some of these infections might actually be caused by a different species is important and will contribute to refine hospital monitoring of Gram-negative infections.

## Results & Discussion

In our lab, species identification of cultured isolates is routinely done by MALDI-ToF MS. All four cases were discovered by partial 16S rRNA sequencing of isolates that obtained an unusually poor (although still valid) score for *E. coli* by MALDI-ToF MS (range 1.9-2.1, versus normally around 2.3). The sequences shared only 98,8% homology with *E. coli*, but was identified as *E. marmotae* with 100% identity. The observation period was from January to July 2021.

### Case reports: *E. marmotae* as cause of septicaemia, cholangitis, pyelonephritis and spondylodiscitis

Case 1: A male in his 80s with haematologic cancer and associated immune deficiency was diagnosed with thoracic spondylodiscitis. A CT-guided aspiration from this focus provided several milliliters of purulent material from which *E. marmotae* was recovered in pure culture (isolate HUSEmarmC1). During the stay, he also had an episode with *Staphylococcus aureus* bacteremia, but *S. aureus* was not detected in the sample from the spondylodiscitis, neither by culture nor by a *S. aureus* specific PCR. The spondylodiscitis was successfully treated with a third generation cephalosporin followed by a course of per oral ciprofloxacin.

Case 2: Pyelonephritis. *E. marmotae* found in pure culture >10^5^ CFU/ml in urine (isolate HUSEmarmC2). No further clinical information permitted.

Case 3: A previously healthy male in his 80s admitted with acute sepsis of unknown origin. In the days prior to this, he had been fertilizing fruit trees with sheep manure. *E. marmotae* was recovered in two sets of blood cultures (isolate HUSEmarmC3). The patient developed a septic shock with multiple organ failure and was transferred to the intensive care unit while treated with a third generation cephalosporin. He recovered well and was dismissed from the hospital to a short term care facility.

Case 4: A male in his 60s with a history of gallstone disease and a well regulated diabetes type II, who developed acute postoperative sepsis with multiple organ failure after cholecystectomy a few days earlier. *E. marmotae* was recovered as the sole agent from two sets of blood culture bottles collected upon admission (isolate HUSEmarmC4a). In addition, *E. marmotae* (isolate HUSEmarmC4b) together with *Streptococcus parasanguinis* and *Enterococcus faecium* were cultured from pus collected from the bile ducts through endoscopic retrograde cholangiopancreatography. The patient responded well to intravenous piperacillin-tazobactam followed by a course of oral trimethoprim-sulfamethoxazole after discharge.

### Whole genome comparisons shows that *E. marmotae* is monophyletic

Whole genome sequencing of the five clinical isolates were performed as described in the Methods chapter. In order to obtain a representative phylogenetic overview of the *Escherichia* genus, we downloaded 36 additional genomes representing *E. marmotae* together with whole genomes representing the main clades, as shown in Denamur et al. (6). Genome accessions as well as relevant metadata is available in supplementary table 1. All genomes were clustered based on pairwise distance using MASH (7). The resulting clusterogram agrees well with established clade designations, and shows that available *E. marmotae* strains constitute a distinct species in the *Escherichia* genus (Figure 1). Other former cryptic *Escherichia* clades recently defined as novel species also cluster by themselves, whereas a selection of diverse *E. coli* appear together, albeit with a larger MASH distance distribution than is the case with *E. marmotae*. The discrepancy is likely a result of strain selection bias due to a larger available pool of *E. coli* sequences to pick from, as well as an historically wide definition of what constitutes *E. coli*.

**Figure 1:**
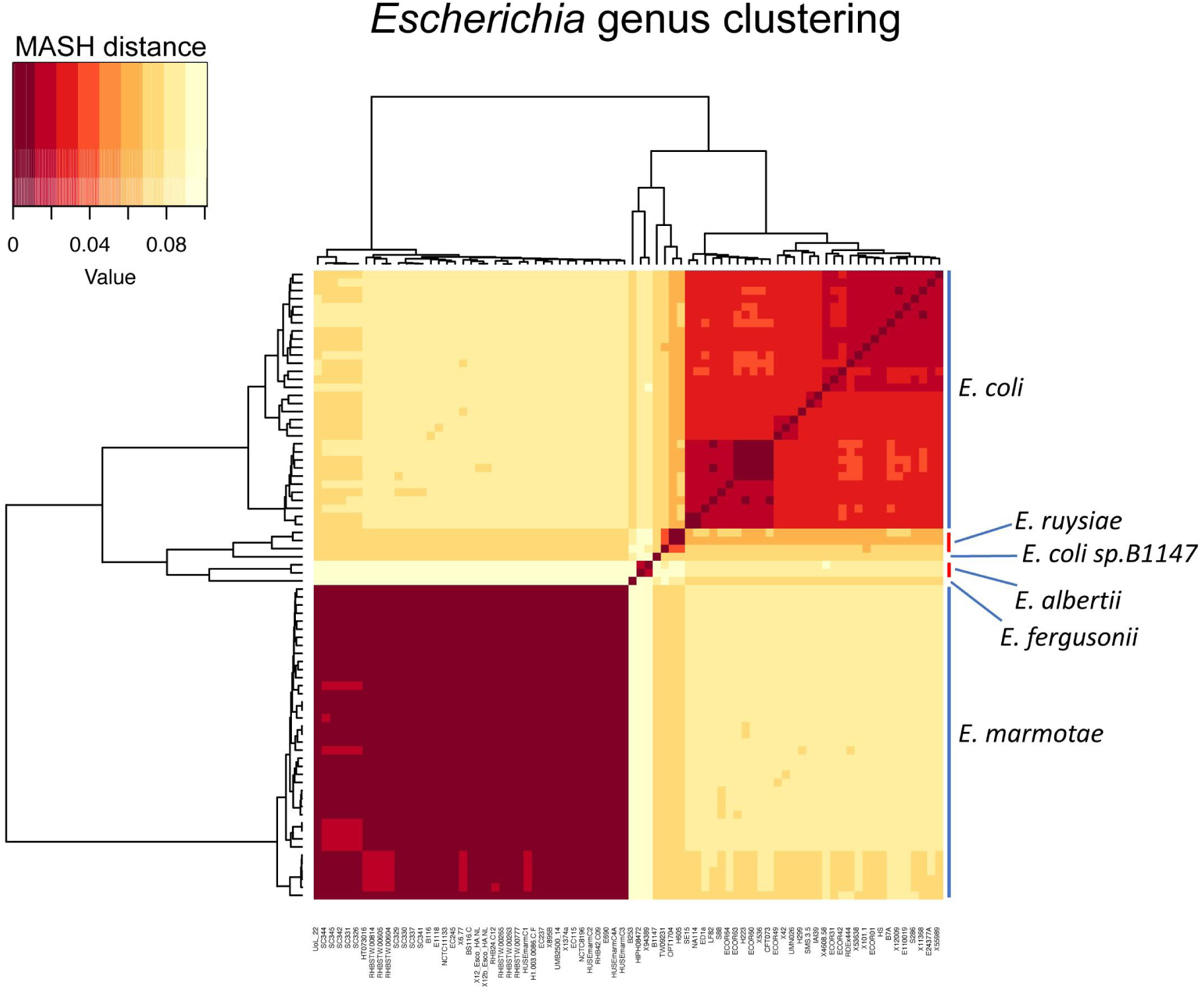
Clustering of Escherichia strains based on small MASH distance (high genetic similarity). Two identical clusterograms surround a heatmap showing pair-to-pair comparisons of all Escherichia strains, with dark red denoting close pairwise similarity. Apparent blocks represent clusters which correspond to species, as described to the right. Strain names are given at the bottom.

### Human clinical strains are mostly scattered within the *E. marmotae* phylogeny

To further investigate the phylogenetic relationships among the 41 *E. marmotae* genomes (36 from GenBank and five from this study) we constructed a separate phylogeny with parsnp (8). The phylogeny was associated with strain origin and presence of antimicrobial resistance genes. We also performed a Roary (9) pan-genome analysis. Six of the available *E. marmotae* whole genomes turned out to represent human isolates (four from feces, one from blood and one from urine, see Fig. 2), originally submitted as *E. coli* but later reassigned as *E. marmotae* by NCBI.

**Figure 2:**
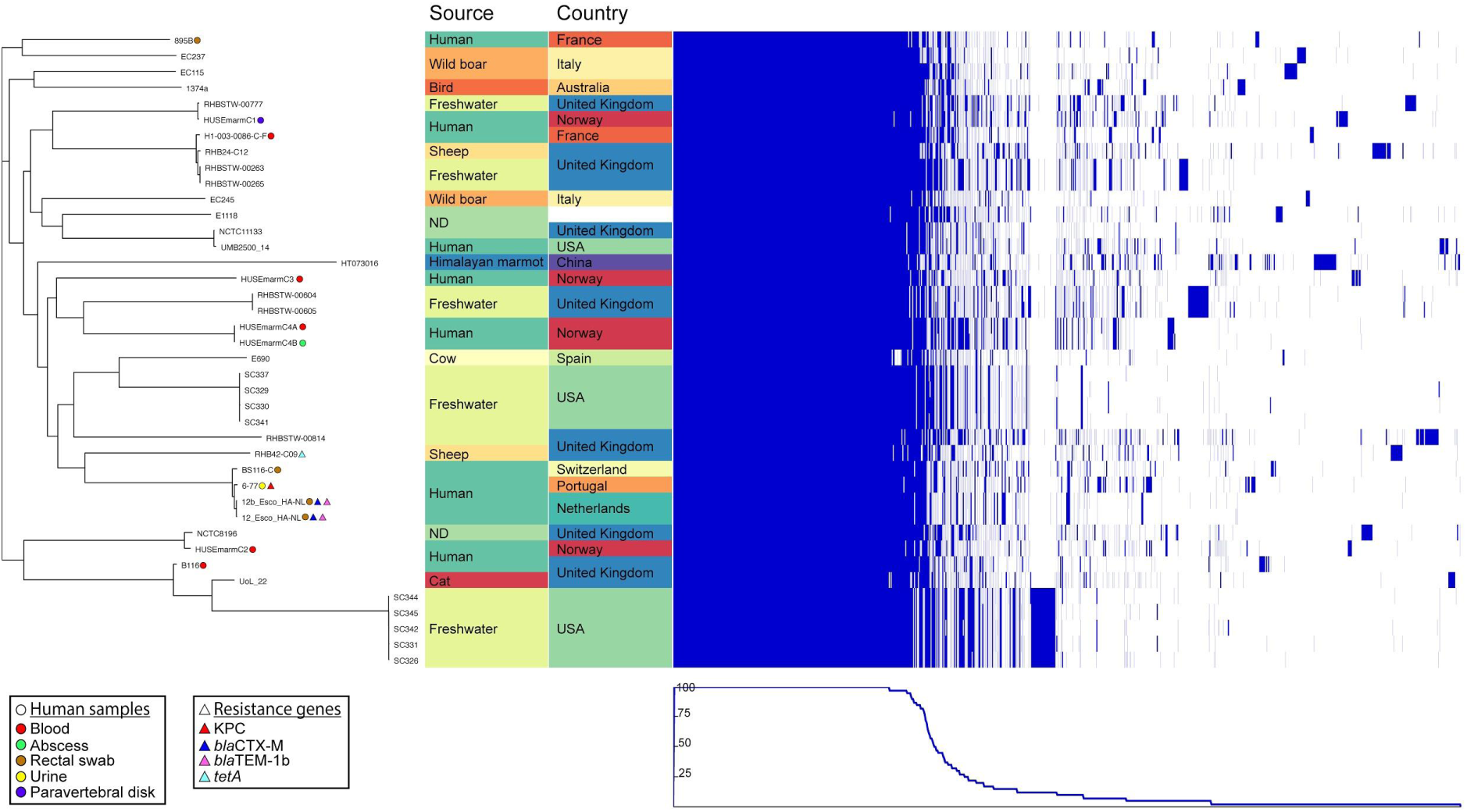
To the left a midpoint rooted phylogeny of E. marmotae strains with highlighting of human samples and isolation sites with coloured dots, and presence of resistance genes with coloured triangles. In the middle, isolate origin by location and country is given and color coded, and rightmost is a pan-genome map showing approximate relative size of the core- and accessory genomes where a blue bar represents presence of a gene. The graph in the bottom right depicts the proportion of isolates harbouring a given gene.

The pan genome of these 41 isolates included 11549 genes, of which 3163 (27,4%) were conserved in all strains. As these few strains were found within a broad context of environments and contained a large accessory genome, we predict *E. marmotae* to be a generalist species like *E. coli*, with similar species ubiquity and similar core- and pan genome features (10).

The 11 genomes from isolates of human origin were scattered throughout the phylogenetic tree, among strains from environmental origins. In two cases, isolates linked to the same patient naturally appeared together, specifically HUSEmarmC4A and B, and presumably 12- and 12b-Esco-HA-NL.

### Available data shows infrequent occurrences of antimicrobial resistance

We could not identify any resistance genes in our five isolates, and all were phenotypically susceptible to the antimicrobials in our standard susceptibility panels (Suppl. table 2).

Among the other *E. marmotae* strains included in this study, genetic resistance markers as determined by the AMRfinder+ database were identified in only four. Among these, three were isolated from human sources thereby linking antimicrobial resistance in this species to human hosts. A single isolate (6-77, UTI, portugal) harboured the *bla*KPC carbapenemase, two isolates, presumably representing the same strain (12-Esco-HA-NL and 12b-Esco-HA-NL, rectal swabs, the Netherlands) had a broad-spectrum beta-lactamase (*bla*CTX-M) in addition to *bla*TEM-1b, and a single strain (RHB43-C09) harboured a gene conferring tetracycline resistance (*tetA*). Only the latter strain did not originate from a human source, but from sheep. The *tetA* gene has previously been found in over 70% of *E. coli* sampled from ruminants (11).

The presence of beta-lactam resistance genes in strains from human sources may represent introduction by horizontal gene transfer within the human gut, as *E. marmotae* is shown to harbour a large accessory genome reminiscent of *E. coli* (Fig. 2, right), and mobile genetic element composition and gene content is visually correlated to local ecology.

Others have found presence of diverse beta lactamases in *E. marmotae* including ESBL and ESBL_CARBA_, and additionally found extensive sharing of resistance genes between different *Enterobacteriaceae* retrieved from the same healthy host faecal samples, indicating frequent spatiotemporal sharing of mobile genetic elements containing resistance genes (12).

### *E. marmotae* virulence re-evaluated

The virulence traits of *E. marmotae* have already been genotypically and phenotypically characterized by Liu *et al*. (2). *However, their analysis included exclusively isolates from the intestine of presumably healthy Marmota himalayana*. Using the vfdb database combined with isolates from a variety of isolation sources, we obtain a broader and somewhat different genetic picture. All genes described here are found with >98% homology to the database entry, as reported by Abricate. In concurrence with Liu *et al*., we find that all *E. marmotae* contain the enterotoxin-encoding gene *astA (13)*. However, only the type strain (HT073016), the sole isolate from *M. himalayana* in our genome collection, contains the *efa1* gene encoding an adherence factor, the *Yersinia* T3SS effector *yopJ*, and a truncated version of the *Shigella*-associated virulence gene *ipaH*.

In addition to the virulence genes previously reported, we find several iron acquisition systems within all *E. marmotae* genomes, notably *entA/B/C/D/E/F/S* encoding the iron siderophore enterobactin (14) and *fepA/B/C/D/G* and *fes* which encodes another siderophore ferrienterochelin (15). All strains also encode *chuS/T/U/V/W/X* involved in heme uptake and metabolism (16, 17). These systems aid in survival during invasive infections by avoiding nutritional immunity, where host sequestration of essential nutrients prevent bacterial survival (16). They are also associated with increased virulence (18).

Further, all *E. marmotae* contain *csgB/D/E/F/G* which are genes facilitating production of extracellular Curli proteins involved in matrixes promoting cellular adhesion and biofilm formation (19), as well as *fimA/B/C/D/E/F/G/H/I* encoding Type 1 fimbriae associated with urinary tract infection (20). They also harbour *ompA*, which encodes the outer membrane protein A associated with several pathogenetic properties including evasion of host defence systems during intra-cellular migration in neonatal meningitis, and inactivation of the complement system (21).

All *E. marmotae* also have a Type 2 Secretion system associated with extracellular release of the enterotoxin produced by ETEC (*gspC/D/E/F/G/H/I/J/K/L/M*) (22). The toxin itself was not found. All also encode *kpsD* which mediates export of group 2 capsular polysaccharides across the outer membrane, notably the O-antigen involved in bacterial survival in blood and urinary tract persistence (23). We also find *kpsM*, associated with the virulence of extraintestinal pathogenic *E. coli* (ExPEC) (24).

Finally, 11 of 41 isolates, with no particular habitat affinity, harbour *faeC/D/E/F/H/I/J* genes which constitute an operon producing fimbriae associated with the pathogenetic processes in enterotoxigenic *E. coli* (ETEC) (25).

By using the ECOH *E. coli in silico* serotyper (26) with abricate, we find all *E. marmotae* strains to contain the *fliC*-H56 flagellar antigen, but a wide array of 12 O-antigens. The H56 serotype is therefore a potential marker of *E. marmotae* in historic data. Interestingly, the H56:O103 serotype of *E. coli* has earlier been reported to populate the intestines of norwegian sheep (27). HUSEmarmC2, the isolate giving pyelonephritis in one patient, was of this serotype. HUSEmarmC3, the isolate retrieved from the patient in previous contact with sheep manure, had the serotype Onovel21:H56.

### Routine identification of *E. marmotae* in the clinical lab

Although we did not systematically search for *E. marmotae*, the four cases from our lab were discovered over a period of only six months. We therefore anticipate that unlike the recently characterized species *E. ruysiae* (Former Escherichia clades III and IV)(25), *E. albertii* and *E. fergusonii* (suppl. material in (6)), *E. marmotae* could represent a relatively regular cause of human invasive infections. Reliable routine identification of *E. marmotae* by clinical laboratories will be necessary to assess the prevalence of infections in humans.

*E. marmotae* or “*Escherichia* cryptic clade V” is phenotypically indistinguishable from *E. coli*. This phenotypic homology is the historic background for the term “cryptic” *Escherichia*. The four clinical cases from our lab were discovered by experienced microbiologists reacting to an unusually low score for *E. coli* by MALDI-ToF MS. At that time, no spectra for *E. marmotae* was available in our MALDI-ToF MS database. This underscores the risk of species misclassification by MALDI-Tof MS when the correct species is not represented in the database, as previously addressed by Body et al (24). They found that *Mycobacterium phocaicum* was regularly misidentified as the closely related *Mycobacterium mucogenicum* due to the lack of spectra for *M. phocaicum*. Unfortunately, the inclusion of a single spectrum for *E. marmotae* (strain HWL_017a HWH) in the latest update of the Bruker Biotyper database (L2020 9607MSP) does not seem to enable robust identification and discrimination from *E. coli*. By re-analysis using the novel database, our isolates obtained scores in the range 1.79 to 2.02 against *E. marmotae* versus 1.70 to 2.06 against *E. coli* (Suppl. table 3). As for *E. coli*, we assume it will be necessary with a wider range of spectra to cover the intra-species variability of *E. marmotae*. Hopefully, the acknowledgement of *E. marmotae* as a human pathogen can stimulate the inclusion of more spectra in MALDI-ToF MS databases.

Available variants of the *E. marmotae* complete 16S rRNA gene display pairwise homologies between 99.4-100% and form a distinct branch in phylogenetic comparisons with the other members of the *Escherichia* genus. Reliable discrimination from *E. coli* based on the 16S rRNA V1-V3 variable areas as commonly used in diagnostic laboratories is formally possible with pairwise homologies varying from 96.6 to 99.0%. Corresponding homologies with *E. albertii, E. fergusonii* and *E. ruysiae* were 96.9-98.6%, 98.0-98.6% and 97.9-99.0% respectively. There appears to have been a lack of 16S rRNA references for the previous *Escherichia* cryptic clade V in GenBank prior to the description of *E. marmotae*. This might have delayed the recognition of *E. marmotae*/*Escherichia* cryptic clade V as a human pathogen.

## Conclusions

*Escherichia marmotae* is a recently described species phenotypically indistinguishable from *E. coli*. We have shown that it can also cause the same types of invasive human infections, involving blood, bone, the urinary tract and the bile duct system, as well as septic shock. It further has the capacity to acquire broad-spectrum β-lactamases including carbapenamases.

A wide range of isolation sources combined with a large accessory genome is indicative of a true generalist bacterium. The extent of encountered virulence genes support that *E. marmotae* has not been acknowledged as a pathogen due to the lack of routine species identification methods rather than recent emergence. The disease-causing strains were scattered among environmental ones, indicating an inherent pathogenic potential in environmental isolates. MALDI-ToF MS has the potential to provide robust routine identification, but a wider range of database *E. marmotae* spectra are needed.

## Methods

### Ethical statement

The study was approved by the Regional ethical committee of Western Norway (REK 322324). Written, informed consent to use clinical data was obtained from three out of four patients. For the fourth patient only microbiological data is provided. All authors declare no conflicts of interest.

### MALDI-ToF MS

For Matrix-assisted laser desorption ionization–time of flight mass spectrometry (MALDI-ToF MS) we used a Microflex LT mass spectrometer (Bruker Daltonics, Billerica, Massachusetts, USA) with database versions K2019 8468MSP (Without *E. marmotae* spectra) during the strain collection period, and version L 2020 9607MSP (Which includes a single *E. marmotae* spectrum) from september 2021. Isolates were smeared directly to the MALDI-ToF MS target plate, and analysed according to the standard routine protocol without an on-plate formic acid treatment.

### DNA extraction

Genomic DNA for Sanger sequencing of the partial 16S rRNA gene and Illumina whole genome sequencing was extracted from overnight bacterial colonies. Mechanical lysis of bacterial cells was performed using the Septifast Lys Kit (Roche Diagnostics, Mannheim, Germany), MagNa Pure Bacteria Lysis Buffer (Roche) and the MagNA Lyser instrument (Roche), followed by extraction on a MagNaPure Compact instrument (Roche) using the MagNA Pure Compact Nucleic Acid isolation Kit I (Roche) according to the manufacturer’s instructions.

### Sanger sequencing of the 16S rRNA gene

Amplification of the variable areas V1-V3 of the 16S bacterial rRNA gene was done using dual priming oligonucleotides as previously described (28), with 5-end modifications as outlined by Dyrhovden et. al. (29) (16S_DPO_Short-F: 5’-AGAGTTTGATCMTGGCTCAIIIIIAACGCT-3’ and 16S_DPO_Short-R: 5’-CGGCTGCTGGCAIIIAITTRGC-3’). The PCR mixture consisted of 12.5 µl of ExTaq SYBR Master Mix (TaKaRa, Otsu, Japan), 0.4 µM of each primer, 8.5 µl of PCR-grade water, and 2 µl of template DNA. The PCR was run on a QuantStudio5 real-time PCR instrument (ThermoFisher, Waltham, MA USA). The PCR thermal profile included an initial polymerase activation step of 10 seconds at 95°C, followed by 45 cycles of 10s at 95°C (melt),15s at 60°C (annealing), and 20s at 72°C (extension). Sanger sequencing was performed in a core facility using an ABI 3730 DNA analyzer (Applied Biosystems/ThermoFisher).

### Whole genome sequencing and bioinformatic analyses

Five isolates from four patients were sequenced on the Illumina MiSeq System with 150 bp paired-end reads. Library was generated using the Nextera XT DNA Library Preparation Kit (Illumina, San Diego, CA, USA). Raw data were deposited in ENA with the accession numbers stated in suppl. table 1. Available genomes representing the *Escherichia* genus and used in the phylogeny published by Denamur *et al*. (6) were downloaded as comparators, see suppl. table 1. Bioinformatic analyses were done by installing and running the Bactopia v.1.7.1 (30) environment, which contains the software and version documentation described below. Briefly, assembly was done with SKESA (31) and annotated with prokka v. 1.14 (32). Using abricate (33), antimicrobial resistance genes were predicted with AMRfinder+ (34), virulence genes predicted with abricate using the VFDB database (35), and serotyping was done using the ECOH database (26). The *Escherichia* genus was clustered by importing the pairwise comparison matrix from MASH (7) into R to create a heatmap using a modified version of the script provided by Desmet et al. (36). A pan-genome analysis of *E. marmotae* was done with Roary (9), and a phylogeny was created with parsnp (8) using -xc flags and using the typestrain HT073016 as reference. Phylogeny with metadata and a pan-genome gene presence/absence scheme was presented via phandango (37). All software was run with standard settings unless otherwise noted.

## Data Availability

All data produced in the present work are contained in the manuscript

## Data availability

The raw data and assemblies of the five sequenced isolates have been published under BioProject PRJEB47670, and BioSample and accession numbers are provided in suppl. table 1.

## Notes

### Competing Interest Statement

The authors have declared no competing interest.

### Funding Statement

This study did not receive any funding

### Author Declarations

The study was approved by the Regional ethical committee of Western Norway (REK 322324). Written, informed consent to use clinical data was obtained from three out of four patients. For the fourth patient only microbiological data is provided.

## References

1. Liu S, Jin D, Lan R, Wang Y, Meng Q, Dai H, Lu S, Hu S, Xu J. 2015. Escherichia marmotae sp. nov., isolated from faeces of Marmota himalayana. Int J Syst Evol Micr 65:2130–2134.

2. Liu S, Feng J, Pu J, Xu X, Lu S, Yang J, Wang Y, Jin D, Du X, Meng X, Luo X, Sun H, Xiong Y, Ye C, Lan R, Xu J. 2019. Genomic and molecular characterisation of Escherichia marmotae from wild rodents in Qinghai-Tibet plateau as a potential pathogen. Sci Rep-uk 9:10619.

3. Lu S, Jin D, Wu S, Yang J, Lan R, Bai X, Liu S, Meng Q, Yuan X, Zhou J, Pu J, Chen Q, Dai H, Hu Y, Xiong Y, Ye C, Xu J. 2016. Insights into the evolution of pathogenicity of Escherichia coli from genomic analysis of intestinal E. coli of Marmota himalayana in Qinghai–Tibet plateau of China. Emerg Microbes Infec 5:1–9.

4. Ocejo M, Tello M, Oporto B, Lavín JL, Hurtado A. 2020. Draft Genome Sequence of Escherichia marmotae E690, Isolated from Beef Cattle. Microbiol Resour Announc 9.

5. Gilroy R, Ravi A, Getino M, Pursley I, Horton DL, Alikhan N-F, Baker D, Gharbi K, Hall N, Watson M, Adriaenssens EM, Foster-Nyarko E, Jarju S, Secka A, Antonio M, Oren A, Chaudhuri RR, Ragione RL, Hildebrand F, Pallen MJ. 2021. Extensive microbial diversity within the chicken gut microbiome revealed by metagenomics and culture. Peerj 9:e10941.

6. Denamur E, Clermont O, Bonacorsi S, Gordon D. 2021. The population genetics of pathogenic Escherichia coli. Nat Rev Microbiol 19:37–54.

7. Ondov BD, Treangen TJ, Melsted P, Mallonee AB, Bergman NH, Koren S, Phillippy AM. 2016. Mash: fast genome and metagenome distance estimation using MinHash. Genome Biol 17:132.

8. Treangen TJ, Ondov BD, Koren S, Phillippy AM. 2014. The Harvest suite for rapid core-genome alignment and visualization of thousands of intraspecific microbial genomes. Genome Biol 15:524.

9. Page AJ, Cummins CA, Hunt M, Wong VK, Reuter S, Holden MTG, Fookes M, Falush D, Keane JA, Parkhill J. 2015. Roary: rapid large-scale prokaryote pan genome analysis. Bioinformatics 31:3691–3693.

10. Touchon M, Perrin A, Sousa JAM de, Vangchhia B, Burn S, O’Brien CL, Denamur E, Gordon D, Rocha EP. 2020. Phylogenetic background and habitat drive the genetic diversification of Escherichia coli. Plos Genet 16:e1008866.

11. Medina A, Horcajo P, Jurado S, Fuente RDL, Ruiz-Santa-Quiteria JA, Domínguez-Bernal G, Orden JA. 2011. Phenotypic and Genotypic Characterization of Antimicrobial Resistance in Enterohemorrhagic Escherichia Coli and Atypical Enteropathogenic E. Coli Strains from Ruminants. J Vet Diagn Invest 23:91–95.

12. Rubin J, Mussio K, Xu Y, Suh J, Riley LW. 2019. Prevalence of antimicrobial resistance genes and integrons in commensal Gram-negative bacteria in a college community. Biorxiv 683524.

13. Maluta RP, Leite JL, Rojas TCG, Scaletsky ICA, Guastalli EAL, Ramos M de C, Silveira WD da. 2016. Variants of ast A gene among extra-intestinal Escherichia coli of human and avian origin. Fems Microbiol Lett fnw285.

14. Raymond KN, Dertz EA, Kim SS. 2003. Enterobactin: An archetype for microbial iron transport. Proc National Acad Sci 100:3584–3588.

15. Schubert S, Fischer D, Heesemann J. 1999. Ferric Enterochelin Transport in Yersinia enterocolitica : Molecular and Evolutionary Aspects. J Bacteriol 181:6387–6395.

16. Richard KL, Kelley BR, Johnson JG. 2019. Heme Uptake and Utilization by Gram-Negative Bacterial Pathogens. Front Cell Infect Mi 9:81.

17. Suits MDL, Lang J, Pal GP, Couture M, Jia Z. 2009. Structure and heme binding properties of Escherichia coli O157:H7 ChuX. Protein Sci 18:825–838.

18. Galardini M, Clermont O, Baron A, Busby B, Dion S, Schubert S, Beltrao P, Denamur E. 2020. Major role of iron uptake systems in the intrinsic extra-intestinal virulence of the genus Escherichia revealed by a genome-wide association study. Plos Genet 16:e1009065.

19. Barnhart MM, Chapman MR. 2006. Curli Biogenesis and Function. Annu Rev Microbiol 60:131–147.

20. Stærk K, Grønnemose RB, Nielsen TK, Petersen NA, Palarasah Y, Torres-Puig S, Møller-Jensen J, Kolmos HJ, Lund L, Andersen TE. 2021. Escherichia coli type-1 fimbriae are critical to overcome initial bottlenecks of infection upon low-dose inoculation in a porcine model of cystitis. Microbiology+ 167.

21. Krishnan S, Prasadarao NV. 2012. Outer membrane protein A and OprF: versatile roles in Gram-negative bacterial infections. Febs J 279:919–931.

22. Strozen TG, Li G, Howard SP. 2012. YghG (GspSβ) Is a Novel Pilot Protein Required for Localization of the GspSβ Type II Secretion System Secretin of Enterotoxigenic Escherichia coli. Infect Immun 80:2608–2622.

23. Sarkar S, Ulett GC, Totsika M, Phan M-D, Schembri MA. 2014. Role of Capsule and O Antigen in the Virulence of Uropathogenic Escherichia coli. Plos One 9:e94786.

24. Zong B, Liu W, Zhang Y, Wang X, Chen H, Tan C. 2016. Effect of kpsM on the virulence of porcine extraintestinal pathogenic Escherichia coli. Fems Microbiol Lett 363:fnw232.

25. Yu M, Qi R, Chen C, Yin J, Ma S, Shi W, Wu Y, Ge J, Jiang Y, Tang L, Xu Y, Li Y. 2017. Immunogenicity of recombinant Lactobacillus casei-expressing F4 (K88) fimbrial adhesin FaeG in conjunction with a heat-labile enterotoxin A (LTAK63) and heat-labile enterotoxin B (LTB) of enterotoxigenic Escherichia coli as an oral adjuvant in mice. J Appl Microbiol 122:506–515.

26. Ingle DJ, Valcanis M, Kuzevski A, Tauschek M, Inouye M, Stinear T, Levine MM, Robins-Browne RM, Holt KE. 2016. In silico serotyping of E. coli from short read data identifies limited novel O-loci but extensive diversity of O:H serotype combinations within and between pathogenic lineages. Microb Genom 2:e000064.

27. Sekse C, Sunde M, Hopp P, Bruheim T, Cudjoe KS, Kvitle B, Urdahl AM. 2013. Occurrence of Potentially Human-Pathogenic Escherichia coli O103 in Norwegian Sheep. Appl Environ Microb 79:7502–7509.

28. Kommedal Ø, Simmon K, Karaca D, Langeland N, Wiker HG. 2012. Dual Priming Oligonucleotides for Broad-Range Amplification of the Bacterial 16S rRNA Gene Directly from Human Clinical Specimens. J Clin Microbiol 50:1289–1294.

29. Dyrhovden R, Nygaard RM, Patel R, Ulvestad E, Kommedal Ø. 2019. The bacterial aetiology of pleural empyema. A descriptive and comparative metagenomic study. Clin Microbiol Infec 25:981–986.

30. Petit RA, Read TD. 2020. Bactopia: a Flexible Pipeline for Complete Analysis of Bacterial Genomes. Msystems 5:e00190–20.

31. Souvorov A, Agarwala R, Lipman DJ. 2018. SKESA: strategic k-mer extension for scrupulous assemblies. Genome Biol 19:153.

32. Seemann T. 2014. Prokka: rapid prokaryotic genome annotation. Bioinformatics 30:2068–2069.

33. Seemann T. ABRicate. https://github.com/tseemann/abricate

34. Feldgarden M, Brover V, Haft DH, Prasad AB, Slotta DJ, Tolstoy I, Tyson GH, Zhao S, Hsu C-H, McDermott PF, Tadesse DA, Morales C, Simmons M, Tillman G, Wasilenko J, Folster JP, Klimke W. 2019. Validating the AMRFinder Tool and Resistance Gene Database by Using Antimicrobial Resistance Genotype-Phenotype Correlations in a Collection of Isolates. Antimicrob Agents Ch 63.

35. Liu B, Zheng D, Jin Q, Chen L, Yang J. 2018. VFDB 2019: a comparative pathogenomic platform with an interactive web interface. Nucleic Acids Res 47:gky1080..

36. Desmet S, Keyser ED, Vaerenbergh JV, Baeyen S, Huylenbroeck JV, Geelen D, Dhooghe E. 2019. Differential efficiency of wild type rhizogenic strains for rol gene transformation of plants. Appl Microbiol Biot 103:6657–6672.

37. Hadfield J, Croucher NJ, Goater RJ, Abudahab K, Aanensen DM, Harris SR. 2017. Phandango: an interactive viewer for bacterial population genomics. Bioinformatics 34:292–293.

